# Imbalance-Aware Robust Representation Learning for Medical Image Binary Classification

**DOI:** 10.64898/2026.08.02.26359489

**Authors:** Manyuan Cheng, Cuiyin Liu, Lei Gu

## Abstract

Class imbalance is a prevalent issue in medical image classification that significantly degrades a model’s capacity to recognize minority-class lesions, thereby restricting its applicability in real-world clinical screening scenarios. Existing studies typically address this problem through data resampling, loss re-weighting, or decision boundary adjustment strategies; however, these methods predominantly focus on compensation during the classification stage. In contrast, the representation learning process in earlier stages is often dominated by easy majority-class samples, and its impact on the feature quality of minority classes has not received adequate attention.

To address this issue, we propose an Imbalance-Aware Robust Representation Learning (IRRL) framework for class-imbalanced medical image classification. IRRL prioritizes the refinement of minority-class-related local representations before global classification. Specifically, implicit local token representations are constructed from convolutional feature maps based on their receptive-field structure. Semantic confidence-guided reliability estimation, difficulty-adaptive supervised contrastive learning, and minority-class prototype regularization are then introduced to improve the learning of informative local representations and hard minority-class samples. Finally, a Transformer performs global context modeling for image-level classification.

Experiments on four public datasets, including ISIC 2018, PAD-UFES-20, OCTID, and BUSI, show that IRRL achieves balanced classification performance, with favorable F1-score and Matthews Correlation Coefficient (MCC) results that reflect improved minority-class recognition quality. The results across datasets with different imaging modalities and imbalance conditions further demonstrate the robustness and consistency of the proposed representation learning strategy.

## 1. Introduction

Skin cancer is one of the most common malignancies globally, with a continuously rising incidence rate. Among these, basal cell carcinoma (BCC) is the most prevalent. Although BCC presents a relatively low risk of distant metastasis, failure to accurately identify it at an early stage can still result in severe local tissue destruction, thereby complicating treatment and increasing the medical burden. Therefore, developing automated skin lesion recognition models applicable to real-world clinical scenarios is of profound clinical significance [1].

To propel research in this domain, scholars have established various benchmarks for skin lesion analysis [2]. In recent years, deep learning has significantly advanced the development of medical image classification. Convolutional Neural Networks (CNNs) are particularly effective at modeling local textures and morphological structures, whereas Vision Transformers (ViTs) exhibit formidable capabilities in global context modeling. Existing studies demonstrate that hybrid architectures combining CNNs and Transformers can jointly facilitate local feature extraction and global semantic modeling, thereby achieving exceptional performance in medical image classification tasks [3–5].

Despite continuous advancements in model architectures, medical image classification still confronts a critical challenge in real-world scenarios: class imbalance. In tasks such as skin lesion recognition, malignant or high-risk lesion samples are typically far fewer than those of common categories, causing the training process to be dominated by majority classes and subsequently impairing the model’s ability to recognize minority classes. Existing methods predominantly address this issue through data resampling, loss re-weighting, or decision boundary adjustment [6–8]. More recent studies have also explored contrastive learning and prototype-based constraints to improve the distribution of minority-class features [9–13]. However, most of these methods focus on classification-level compensation or global representation optimization, paying insufficient attention to the local discriminative information of key lesion regions, the enhancement of hard minority-class samples, and the structural stability of minority-class representations. Particularly in medical images, diagnostically significant lesions often occupy only a marginal portion of the image. If these regions are not effectively highlighted during the representation learning process, minority-class features may be diluted by background information or majority-class patterns, thereby degrading the ultimate recognition performance.

Motivated by the aforementioned analysis, we propose a robust representation learning method for class-imbalanced medical image classification, termed Imbalance-Aware Robust Representation Learning (IRRL). Unlike traditional methods that primarily focus on compensation during the classification stage, IRRL shifts the paradigm to the representation learning stage, emphasizing the enhancement of local representations for key regions and hard minority-class samples prior to global context modeling. Specifically, we construct implicit local token representations based on the receptive field structure of convolutional feature maps, treating each spatial position within the feature map as a local implicit token. This design preserves the advantages of convolutional local structures while maintaining low computational overhead, thereby facilitating the fine-grained modeling of lesion regions. Building upon this foundation, semantic confidence modulation, difficulty-adaptive supervised contrastive learning, and a minority-class prototype constraint are further introduced to amplify the contributions of highly informative regions and hard minority-class samples during training. Ultimately, the optimized local representations are fed into a Transformer for global context modeling to execute image-level classification.

The main contributions of this paper are summarized as follows:

- We propose a robust representation learning framework, IRRL, for class-imbalanced medical image classification. This framework shifts the imbalance mitigation process from the conventional classification stage to the representation learning stage, aiming to enhance the discriminability and stability of minority-class features.
- We develop an imbalance-aware optimization strategy based on implicit local tokens. By leveraging semantic confidence modulation and difficulty-adaptive supervised contrastive learning, this strategy reinforces the representation learning of key lesion regions and hard minority-class samples.
- We design a confidence-guided minority-class prototype regularization method to constrain the minority-class feature distribution and improve its intra-class compactness.
- We conduct extensive experiments on four public medical image datasets, demonstrating the effectiveness of IRRL in minority-class recognition and its robustness across diverse medical imaging scenarios.

### 1.1. CNN–Transformer Representation Learning in Medical Image Classification

Hybrid architectures that integrate the local perception capabilities of Convolutional Neural Networks (CNNs) with the global dependency modeling of Vision Transformers (ViTs) have emerged as one of the mainstream paradigms in medical image analysis [1–5]. However, the effectiveness of the self-attention mechanism heavily relies on the quality of the input token representations. In highly class-imbalanced medical scenarios, majority classes (e.g., backgrounds or common lesions) often dominate the early stages of representation learning. This dominance leads to semantic dilution or noise interference in the extracted local features, thereby restricting the Transformer’s capability to focus on minority-class lesion regions. Furthermore, conventional explicit patch partitioning introduces extra computational overhead and can potentially disrupt the inherent spatial topology of medical images. In contrast, constructing implicit tokens from convolutional feature maps can more naturally preserve the structural information within the receptive field. Based on this observation, we argue that the optimization focus should shift toward the representation learning stage, enhancing the discriminability and robustness of local implicit tokens to provide a more reliable feature foundation for minority-class recognition under class-imbalanced conditions.

### 1.2. Class-Imbalanced Medical Image Classification

Class imbalance is a long-standing challenge in medical image analysis. Existing solutions can be broadly categorized into three types: data-level resampling, loss-level re-weighting (e.g., Focal Loss), and decision boundary optimization (e.g., LDAM) [6–8]. Although these methods partially alleviate the imbalance in class frequencies, they primarily focus on macro-level compensation during the decision stage. Consequently, they often overlook the structural imbalance during the representation learning processwhere the model is highly susceptible to being dominated by a vast number of easy majority-class samples during early optimization, causing the representations of minority-class lesions to be diluted or subsumed within the feature space.

To achieve finer-grained optimization, recent studies have explored sample-level dynamic re-weighting mechanisms. For instance, Ansari et al. [14] proposed dynamically adjusting sample weights through real-time difficulty estimation to mitigate algorithmic bias caused by data imbalance and skin tone variations. This approach demonstrates the potential of difficulty-aware optimization under long-tailed distributions. Nevertheless, such methods still predominantly focus on macro-level bias correction. We argue that to further enhance the discriminability of minority classes, the focus must be shifted from the conventional classification stage to the representation learning stage.

### 1.3. Contrastive Learning and Robust Minority-Class Representation

Contrastive learning facilitates the learning of discriminative features by pulling positive sample pairs closer and pushing negative sample pairs further apart in the embedding space. Under a supervised setting, supervised contrastive learning [9] leverages label information to construct structural constraints, which has become an important paradigm for improving minority-class representations in imbalanced learning tasks. For example, BCL [10] and ProCo [11] improve feature distribution under long-tailed settings through balanced loss weights and prototype-aware mechanisms, respectively. ECL [12] and BPaCo [13] further introduce class augmentation and balanced parameter constraints, significantly enhancing the compactness of minority-class features.

Despite these advancements, two critical limitations remain. The first is the spatial limitation: most contrastive learning methods operate on image-level global representations, ignoring the locality of lesion regions in medical images. In dermoscopy or ultrasound images, if local discriminative information is diluted by background noise, global optimization alone may be insufficient to ensure sensitivity to subtle minority-class lesions. The second is the sample limitation: traditional contrastive learning implicitly assumes that all samples contribute equally to optimization, failing to fully account for the vast disparities in sample difficulty inherent in medical imaging tasks. To address these issues, we propose the IRRL framework. Compared with existing methods, IRRL emphasizes filtering background noise through semantic confidence at the local representation stage and introduces a difficulty-adaptive mechanism to dynamically amplify the contributions of high-risk minority-class samples, thereby achieving more robust representation learning under extreme class-imbalanced conditions.

## 2. Method

### 2.1. Problem Definition and Overall Framework

To address the severe class imbalance challenge in medical image classification, we propose an Imbalance-Aware Robust Representation Learning (IRRL) framework. Unlike traditional methods that primarily rely on re-weighting during the decision stage, IRRL introduces proactive intervention during the representation learning phase. It reshapes the distribution of the learned feature space through a “local-to-global” two-stage optimization paradigm. The overall architecture of IRRL is illustrated in Fig. 1.

**Fig. 1:**
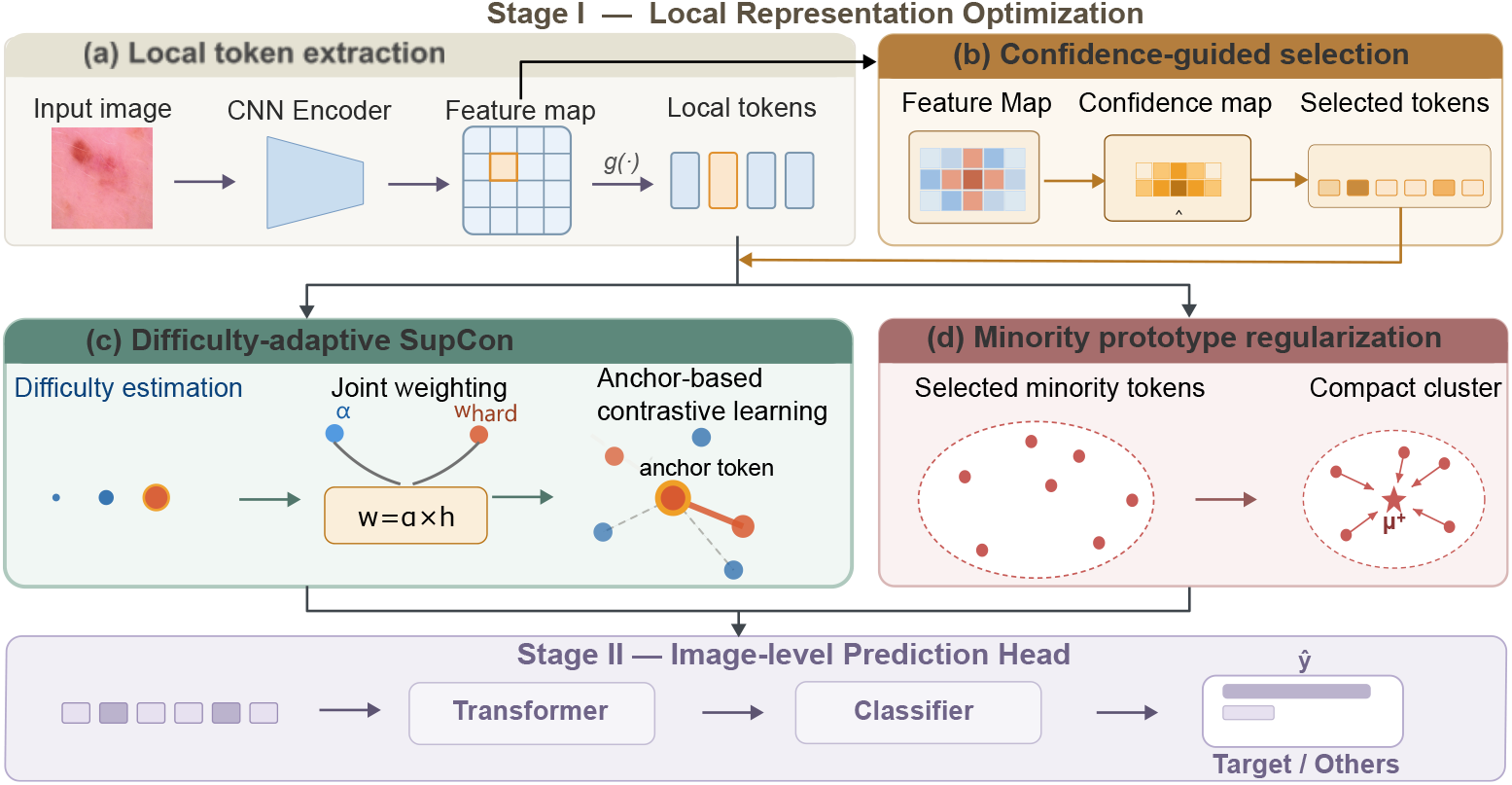
Overall architecture of the IRRL framework. The framework consists of two core stages: (1) The local representation optimization stage, which refines the CNN-derived implicit tokens via semantic confidence weighting, Difficulty-Adaptive Supervised Contrastive learning (DASC), and minority-class prototype regularization (Proto); (2) The global prediction stage, which employs a Transformer to conduct global context modeling on the refined tokens and yields the image-level classification results.

Specifically, the operational workflow of the framework proceeds as follows:

- **Stage I (Local Representation Refinement):** Given an input image, a convolutional encoder first extracts local feature maps. We leverage the receptive field structure to construct implicit token sequences to preserve spatial topological priors, and introduce semantic confidence estimation, difficulty-adaptive weighting, and minority-class prototype regularization. This stage aims to suppress background noise, amplify the supervision signals for hard minority-class samples, and reduce the intra-class variance of minority-class features.
- **Stage II (Global Context Modeling):** The robust local representations optimized in Stage I, integrated with a learnable [CLS] token and positional embeddings, are fed into a Transformer for image-level global context modeling, ultimately outputting the classification prediction.

Through the design of “local refinement followed by global modeling,” this two-stage paradigm mitigates the adverse effects of imbalanced distributions on classification decisions directly at the representation level, thereby providing a more stable feature foundation for subsequent discrimination.

### 2.2. Receptive Field-Based Local Implicit Token Embedding

Let the training dataset be defined as:

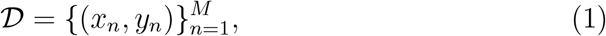

where 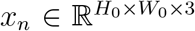 denotes the *n*-th input image, *y*_*n*_ ∈ {0, 1} represents the corresponding binary classification label, and *M* is the total number of training samples.

In Stage I, a convolutional encoder *f* (·) is employed to extract the feature map of the input image:

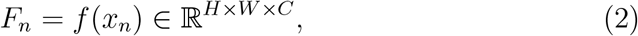

where *C* denotes the number of feature channels, and *H* and *W* represent the height and width of the feature map, respectively. Since each spatial position within the convolutional feature map corresponds to a local receptive field region in the input image, each spatial location can be naturally viewed as a local implicit token.

Specifically, in convolutional networks, each spatial position (*i, j*) in the feature map corresponds to a receptive field region in the input image. Therefore, we regard each position of the feature map as a local implicit token representation and construct the local token embedding as follows:

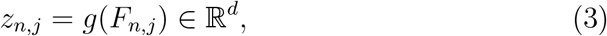

where *F*_*n,j*_ ∈ ℝ^*C*^ denotes the feature vector at the spatial position (*n, j*) of the feature map, *g*(·) is a linear projection head, and *z*_*n,j*_ ∈ ℝ^*d*^ represents the corresponding local implicit token embedding. Subsequently, all local token embeddings are arranged into a token sequence:

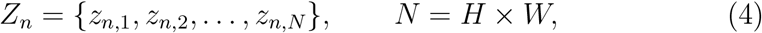

where *N* denotes the total number of local implicit tokens.

This receptive field-based local implicit token construction strategy preserves the local texture information and spatial topology encoded within the convolutional features, while avoiding the extra computational overhead introduced by explicit patch partitioning. Consequently, it provides a more natural and stable input representation for the subsequent local representation refinement and global context modeling.

### 2.3. Semantic Confidence-Guided Token Reliability Estimation

Medical images typically contain extensive non-diagnostic regions, whereas clinically significant lesions often occupy only a limited spatial area. Therefore, under class-imbalanced conditions, if all local tokens are directly optimized with equal contributions, irrelevant background signals might be introduced during representation learning, thereby diminishing the discriminative capability of minority-class-related regions.

Unlike traditional attention mechanisms primarily utilized to recalibrate feature responses, IRRL introduces semantic confidence estimation to evaluate the optimization reliability of local implicit tokens. The estimated confidence scores are not applied to directly modify feature activations; rather, they serve as guiding signals to select highly informative tokens and assign optimization priorities during the representation learning process. Based on the constructed local implicit token sequence *Z*_*n*_, we further estimate the semantic confidence of each token.

IRRL employs two complementary reliability estimation paths to capture the semantic relevance and spatial importance of local tokens, respectively. Specifically, the channel-guided path first acquires channel-wise semantic responses through global context aggregation to measure the contributions of different feature channels to class discrimination. Meanwhile, the spatial-guided path generates spatial reliability scores based on local spatial feature responses to evaluate the likelihood that tokens at varying spatial locations contain valid diagnostic information. Subsequently, these two types of responses are fused and mapped via a Sigmoid function to yield the semantic confidence score for the local token:

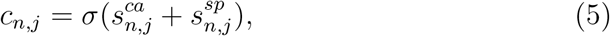

where *c*_*n,j*_ denotes the semantic confidence score of the *j*-th local token in the *n*-th sample, *σ*(·) represents the Sigmoid activation function, and 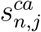 and 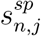 denote the reliability responses generated by the channel-guided path and the spatial-guided path, respectively.

The obtained confidence scores are interpreted as reliability estimates of the tokens, rather than traditional attention coefficients. A higher confidence value indicates that the corresponding token is more likely to contain clinically significant information and should make a greater contribution to the subsequent representation-level optimization.

The confidence-guided token reliability estimation serves as a bridge between local feature extraction and imbalance-aware representation learning. Specifically, the confidence scores are further applied in the proposed difficulty-adaptive contrastive learning and minority-class prototype regularization modules, enabling the model to focus on reliable minority-class-related representations while mitigating interference caused by irrelevant background regions.

### 2.4. Difficulty-Adaptive Supervised Contrastive Representation Learning

Although supervised contrastive learning can enhance feature discriminability by reducing intra-class variance and expanding inter-class margins, traditional contrastive objectives treat all positive and negative sample pairs indiscriminately. Under severe class imbalance, this uniform optimization strategy may be dominated by majority-class samples, paying insufficient attention to hard minority-class-related representations.

To address this limitation, IRRL introduces a difficulty-adaptive supervised contrastive representation learning strategy, which dynamically adjusts the contribution of each local token based on its classification difficulty.

For each local token representation *z*_*n,j*_, the model predicts its binary classification probability distribution:

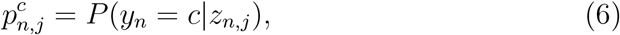

where *c* denotes the class index. The probability corresponding to the ground-truth class is defined as:

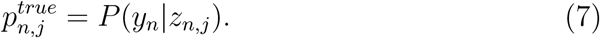

The difficulty coefficient of each token is calculated as follows:

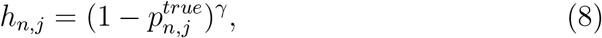

where *γ* controls the degree of focus on hard samples. A larger value of *h*_*n,j*_ indicates greater classification difficulty for the corresponding token, warranting stronger optimization attention.

Unlike Focal Loss, which adjusts the classification loss based on prediction confidence, the proposed difficulty coefficient is integrated into the contrastive representation optimization process. Therefore, IRRL encourages the model to construct more discriminative embeddings for hard minority-class-related tokens, rather than merely correcting classification bias.

The difficulty-adaptive supervised contrastive loss is formulated as:

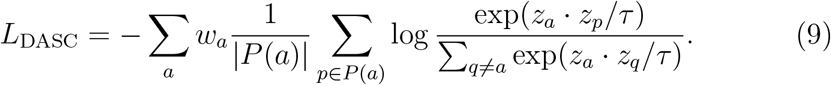

where *a* denotes the anchor token, and *P* (*a*) represents the set of other local tokens within the current mini-batch that share the same image-level class label as the anchor token *a. p* denotes a positive sample token sampled from *P* (*a*), *q* denotes other tokens used for contrastive normalization, and *τ* is a temperature parameter controlling the smoothness of the contrastive distribution. *w*_*a*_ = *α*_*a*_*h*_*a*_ denotes the joint optimization weight for the anchor token *a*, where *α*_*a*_ represents the class imbalance weight and *h*_*a*_ denotes the difficulty coefficient of the corresponding token.

By introducing difficulty-aware optimization into contrastive representation learning, IRRL enables the model to allocate more learning capacity to ambiguous minority-class representations while preserving global feature discriminability.

### 2.5. Confidence-Guided Minority Representation Regularization

Although supervised contrastive learning can improve inter-class separability, minority-class representations may still exhibit substantial intra-class variance due to the limited number of minority-class samples. Furthermore, image-level annotations do not provide explicit local lesion labels; therefore, it is inappropriate to directly designate all local tokens in minority-class images as lesion-related representations.

To address this issue, IRRL introduces a confidence-guided minority representation regularization strategy, which selects reliable minority-class-related local tokens based on image-level labels and semantic confidence scores.

Unlike treating all tokens from minority-class images as positive representations, we exclusively select tokens with high confidence:

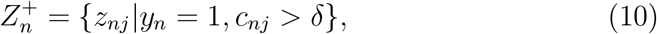

where *δ* is the confidence threshold.

The minority-class prototype is computed utilizing the selected minority-class-related tokens:

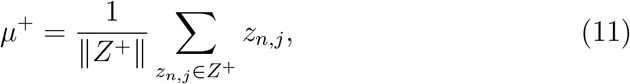

where *Z*^+^ denotes the set of reliable minority-class-related tokens within the current mini-batch.

The prototype regularization objective is formulated as:

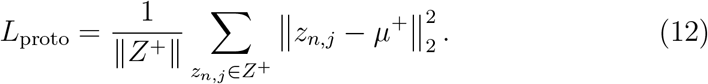

This design avoids introducing background noise into minority-class representation learning and enables the prototype constraint to focus on lesion-related regions enriched with semantic information. Compared to supervised contrastive learning, which primarily enhances inter-class discriminability, the proposed prototype regularization further improves the compactness and stability of minority-class-related representations.

### 2.6. Stage II: Transformer-Based Global Context Modeling

Upon completing the local representation refinement, IRRL feeds the local implicit tokens generated in Stage I into a Transformer for image-level global context modeling. The underlying motivation for this design is that although the self-attention mechanism of Transformers is highly effective in modeling long-range dependencies, its performance is heavily contingent upon the quality of the input tokens. If the input tokens still suffer from substantial background interference or unstable minority-class representations, the efficacy of global modeling could be significantly compromised. Therefore, IRRL adopts a “local refinement followed by global modeling” two-stage strategy to fully harness the complementary advantages of CNNs and Transformers.

Specifically, the local implicit tokens are first mapped from the local representation space to the Transformer latent space via a linear projection:

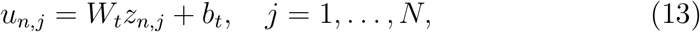

where *W*_*t*_ and *b*_*t*_ denote the learnable projection matrix and bias term, respectively, and *u*_*n,j*_ represents the representation of the *j*-th local token in the *n*-th sample within the Transformer latent space.

Subsequently, the projected local tokens are concatenated with a learnable [CLS] token, and positional encodings are added to construct the input sequence for the Transformer:

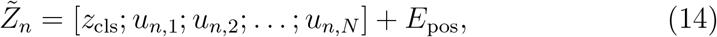

where *z*_cls_ denotes the learnable classification token, and *E*_pos_ represents the positional encoding.

The resulting input sequence is defined as the initial hidden state of the Transformer encoder:

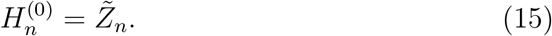

This sequence is then fed into a Vision Transformer consisting of *L* encoder layers, where the hidden representations are progressively updated layer by layer:

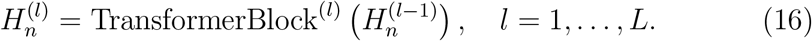

The output corresponding to the [CLS] token is taken as the image-level representation *h*_n,cls_, which is further passed through a classification head to yield the predicted class probability:

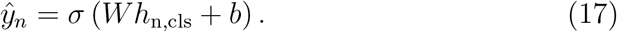

Since the first stage has already performed targeted optimization on the local minority-class representations, the second stage employs the standard binary cross-entropy loss without requiring additional class re-weighting. The corresponding image-level classification loss is defined as:

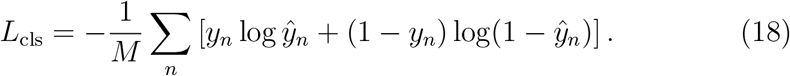

Refraining from any class re-weighting strategies in this stage preserves the stability of the decision-level training.

### 2.7. Overall Optimization Objective

The proposed IRRL framework jointly optimizes classification discriminability and imbalance-aware representation learning. Specifically, the overall objective comprises three complementary components: the classification loss, the difficulty-adaptive supervised contrastive representation loss, and the minority-class representation prototype regularization loss.

The total training objective is formulated as:

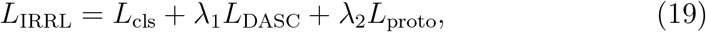

where *L*_cls_ denotes the traditional classification loss, *L*_DASC_ represents the proposed difficulty-adaptive supervised contrastive representation learning objective, and *L*_proto_ stands for the confidence-guided minority representation regularization loss.

The classification loss preserves the model’s discriminative capability for the final class prediction. The difficulty-adaptive contrastive objective focuses on reshaping the feature space by allocating greater optimization emphasis to ambiguous minority-class-related representations. Meanwhile, the prototype regularization term constrains reliable minority-class representations within a compact feature distribution, thereby mitigating the intra-class variance induced by the limited number of minority-class samples.

Through the joint optimization of these three objectives, IRRL shifts the imbalance mitigation process from traditional decision-level adjustment to representation-level refinement, thereby achieving more robust minority-class feature learning in long-tailed medical image scenarios.

Overall, the essence of IRRL lies not in modifying the original sample distribution, but in elevating local representation quality through local semantic modeling, difficulty-adaptive optimization, and minority-class prototype constraints, subsequently achieving robust image-level discrimination via global context modeling. Consequently, IRRL is capable of improving the recognition quality of minority classes in class-imbalanced medical image classification tasks, striking a more optimal balance among Sensitivity, Specificity, and Precision.

## 3. Experiments

### 3.1. Experimental Settings

#### 3.1.1. Datasets and Task Definition

To systematically evaluate the efficacy of the proposed method (IRRL) in class-imbalanced medical image classification, four public datasets encompassing diverse imaging modalities are selected for experimentation. To simulate the clinical screening scenario of “accurately identifying specific high-risk lesions from complex backgrounds,” a unified binary classification setting of “Target vs. Others” is formulated across all datasets. Detailed task definitions and dataset statistics are summarized in Table 1.

**Table 1:** Summary of datasets used in this study, including ISIC 2018, PAD-UFES-20, OCTID, and BUSI. The table reports imaging modalities, task definitions, numbers of minority and majority samples, and imbalance ratios (IR).

| Dataset | Imaging Modality | Task (Minority vs. Majority) | Minority Samples | Majority Samples | Imbalance Ratio (IR) |
| --- | --- | --- | --- | --- | --- |
| ISIC 2018 | Dermoscopic images | BCC vs. Others | 514 | 9,501 | 18.5 : 1 |
| PAD-UFES-20 | Clinical smartphone images | BCC vs. Others | 845 | 1,453 | 1.7 : 1 |
| OCTID | Retinal OCT images | AMD (Age-related Macular Degeneration) vs. Others | 55 | 517 | 9.4 : 1 |
| BUSI | Breast ultrasound images | Malignant vs. Others | 210 | 570 | 2.7 : 1 |

First, the dermoscopy benchmark dataset ISIC 2018 [15] is utilized for the primary experiments, focusing on the identification of basal cell carcinoma (BCC vs. Others). Under this setting, the imbalance ratio reaches up to 18.5:1. Such extreme imbalance often leads to the optimization process being dominated by majority-class samples, making it a challenging benchmark for evaluating the robustness of minority-class representation learning.

Furthermore, PAD-UFES-20 [16], OCTID [17], and BUSI [18] are utilized for supplementary evaluation. Recent studies on medical image benchmarks [2] emphasize the importance of evaluating long-tailed learning methods across multiple modalities and clinical scenarios. On PAD-UFES-20, the identical BCC vs. Others setting is adopted, whereas OCTID and BUSI are formulated as AMD (Age-related Macular Degeneration) vs. Others and Malignant vs. Others classification tasks, respectively.

As illustrated in Table 1, these supplementary datasets cover a diverse array of imaging modalities, including clinical smartphone images, retinal OCT scans, and ultrasound imaging. Moreover, their imbalance ratios (IR) are 1.7, 9.4, and 2.7, respectively. Such a diversified experimental setup, encompassing cross-organ and cross-device variations, enables a comprehensive assessment of the robustness and generalization capability of the proposed method under complex clinical distributions.

#### 3.1.2. Implementation Details

All models are implemented using the PyTorch framework, and all experiments are conducted on a workstation equipped with an NVIDIA GeForce RTX 4070 GPU. All input images are resized to 224 × 224 and normalized before being fed into the network. During training, standard data augmentation strategies are applied, including random horizontal flipping, random vertical flipping, random rotation, color jittering, and random erasing. The batch size is set to 32, and a fixed random seed of 42 is used throughout the experiments to improve reproducibility. The same data partitioning protocol is adopted for all comparison methods to ensure a fair evaluation.

For datasets with predefined official training and test sets, the official training set is further divided into training and validation subsets at a ratio of 8:2 using stratified sampling, while the official test set is retained exclusively for final performance evaluation. For datasets without predefined official splits, the entire dataset is first divided into training and test subsets at a ratio of 8:2 using stratified sampling, after which the resulting training subset is further divided into training and validation subsets at a ratio of 8:2, also using stratified sampling.

Considering the limited number of minority-class samples in some datasets, the validation split is additionally constrained to retain a prespecified minimum number of minority-class samples to improve the stability of validation-based model selection. This constraint is determined solely from the class-label distribution before model training and does not involve any model-performance information.

Regarding the network architecture, ResNet-50 is employed as the convolutional encoder to extract local feature maps, from which receptive-field-based implicit local token representations are constructed. The dimension of the local token projection is set to 128. The refined local representations are subsequently fed into a Transformer for global context modeling. The Transformer embedding dimension is set to 512, with 4 encoder layers and 8 attention heads.

During Stage I, the local representation learning module is trained for a maximum of 50 epochs using the AdamW optimizer. The initial learning rate is set to 1 × 10^−4^, with a weight decay of 5 × 10^−4^ and a 5-epoch warm-up strategy. The temperature parameter *τ* in the supervised contrastive objective is set to 0.07, and the difficulty focusing parameter *γ* is set to 2.0. The loss coefficients *λ*_1_ and *λ*_2_ are set to 0.5 and 0.3, respectively. To reduce the influence of unstable difficulty estimation during the early stage of training, uniform difficulty weighting is adopted during the first 5 epochs before activating the difficulty-adaptive weighting mechanism.

During Stage II, the local representations learned in Stage I are further integrated with Transformer-based global context modeling, and the model is fine-tuned for a maximum of 60 epochs. The Transformer-related components are optimized with a learning rate of 5 × 10^−5^, whereas the convolutional encoder is assigned a smaller learning rate of 5 × 10^−6^ to preserve the previously learned local representations. The convolutional encoder is unfrozen after the first 5 epochs, and a layer-wise learning-rate decay factor of 0.75 is adopted. A 5-epoch warm-up strategy and an early-stopping patience of 15 epochs are used during this stage. Cosine annealing learning-rate scheduling and gradient clipping are employed to improve optimization stability.

For all methods, no dynamic threshold searching or additional threshold adjustment based on the validation set is performed. A fixed decision threshold is used for evaluation, and the model checkpoint achieving the best AUC on the validation set is selected for final testing.

#### 3.1.3. Evaluation Metrics

Considering that accuracy is highly susceptible to the dominance of the majority class under imbalanced settings, thereby failing to adequately reflect the recognition performance of the minority class, we primarily report the F1-score, Matthews Correlation Coefficient (MCC), and Area Under the ROC Curve (AUC). The F1-score evaluates the balance between Precision and Recall, rendering it particularly suitable for assessing minority-class detection performance. Meanwhile, the MCC comprehensively accounts for True Positives (TP), True Negatives (TN), False Positives (FP), and False Negatives (FN), thus providing a more holistic evaluation for class-imbalanced tasks.

Furthermore, we introduce geometric embedding metrics, including intra-class variance, inter-class distance, and the Silhouette Score, to analyze the structural disparities within the learned representation space. These metrics serve exclusively as auxiliary evidence for mechanistic analysis, rather than as primary performance indicators.

### 3.2. Comparison Methods

To comprehensively evaluate the efficacy of the proposed method, four categories of representative baseline methods are introduced for comparison.

The first category comprises Architectural Baselines, including ResNet-50 [4], ViT-B/16 [5], and CNN+Transformer. These are utilized to analyze performance disparities among distinct backbone architectures under the class-imbalanced medical image classification setting, and to ascertain whether the performance gains are merely attributable to backbone upgrades.

The second category encompasses classical imbalanced learning methods, such as Focal Loss [6], Class-Balanced Loss [7], LDAM-DRW [8], and Random OverSampling (ROS). These approaches address class imbalance from the perspectives of loss re-weighting, decision boundary adjustment, and data resampling, respectively.

The third category consists of representation learning and contrastive learning-based methods, such as BCL [10], ProCo [11], ECL [12], and BPaCo [13]. These methods focus on refining the feature space structure under long-tailed distributions via contrastive objectives or representation constraints, making them closely related to the representation-level imbalanced learning strategy proposed in this work.

The fourth category is composed of external frameworks, including MedViT [3] and VGG19-RSPDA [19]. These methods represent strong general-purpose baselines in medical image classification and task-specific hybrid frameworks for skin lesion analysis, respectively. Comparisons against them serve to evaluate whether merely employing stronger backbones or more complex architectures is sufficient to overcome the minority-class recognition challenges posed by class imbalance.

### 3.3. Main Experimental Results on ISIC 2018

Table 2 reports the classification results of different methods on the ISIC 2018 dataset. Overall, the results indicate that under the class-imbalanced setting, the performance improvements yielded by various methods exhibit distinct disparities in their optimization focus. Certain methods achieve higher performance in Sensitivity, thereby enhancing the recall of the minority class, whereas others demonstrate advantages in Specificity or AUC, reflecting stronger stability in discriminating the majority class. However, improvements in a single metric do not necessarily translate into superior overall discriminative performance.

**Table 2:** Presents the performance comparison of different methods on the ISIC 2018 dataset. Metrics include AUC, Sensitivity, Specificity, Accuracy, F1-score, and MCC.

| Method Category | Method | Backbone | AUC | Sens. | Spec. | ACC | F1-score | MCC |
| --- | --- | --- | --- | --- | --- | --- | --- | --- |
| Architectures | ResNet-50 | CNN | 0.9643 | 0.5484 | 0.9915 | 0.9643 | 0.6538 | 0.6492 |
|  | ViT-B/16 | Transformer | 0.9574 | 0.4194 | 0.9866 | 0.9517 | 0.5166 | 0.5078 |
|  | Hybrid (baseline) | Hybrid | 0.9669 | 0.5484 | **0.9944** | 0.9669 | 0.6711 | 0.6734 |
| Hybrid FWs | MedViT | Hybrid | 0.9104 | 0.6559 | 0.9063 | 0.8909 | 0.4251 | 0.4039 |
|  | VGG19-RSPDA | Hybrid | 0.9787 | 0.3763 | 0.9936 | 0.9557 | 0.5109 | 0.5288 |
| Loss Opt. | + Focal Loss | Hybrid | 0.9770 | 0.7312 | 0.9789 | 0.9636 | 0.7120 | 0.6929 |
|  | + CB Loss | Hybrid | 0.9730 | 0.8387 | 0.9514 | 0.9444 | 0.6500 | 0.6407 |
|  | + LDAM-DRW | Hybrid | 0.9697 | 0.6559 | 0.9887 | 0.9683 | 0.7176 | 0.7045 |
|  | + ROS | Hybrid | 0.9700 | 0.7312 | 0.9725 | 0.9577 | 0.6800 | 0.6593 |
| Rep. Learning | ECL | Hybrid | 0.9677 | 0.7957 | 0.9676 | 0.9570 | 0.6948 | 0.6784 |
|  | ProCo | Hybrid | 0.9724 | 0.7742 | 0.9598 | 0.9484 | 0.6486 | 0.6313 |
|  | BCL | Hybrid | 0.9640 | **0.8710** | 0.9563 | 0.9511 | 0.6864 | 0.6792 |
|  | BPaCo | Hybrid | **0.9803** | 0.6237 | 0.9930 | **0.9702** | 0.7205 | 0.7148 |
| Proposed | **IRRL (Ours)** | Hybrid | 0.9702 | 0.7312 | 0.9845 | 0.9689 | **0.7432** | **0.7267** |

In contrast, IRRL achieves the optimal results in the F1-score and Matthews Correlation Coefficient (MCC), two metrics highly representative of overall performance in class-imbalanced scenarios. Specifically, IRRL attains an F1-score of 0.7432 and an MCC of 0.7267, exhibiting a more balanced trade-off within the Precision-Recall space.

The superiority of IRRL is not attributable to the extreme optimization of a single metric, but rather to achieving a more harmonized synergistic improvement across multiple key evaluation criteria. Such balanced performance is particularly crucial for class-imbalanced medical image classification tasks. Overemphasizing recall may introduce a substantial number of false positives, thereby degrading clinical applicability; conversely, an excessive focus on specificity or precision may lead to the missed diagnosis of clinically significant minority-class cases. The results in Table 2 demonstrate that IRRL does not merely improve performance by naively expanding positive predictions, but rather achieves a more controllable trade-off in the Precision-Recall space, thereby yielding higher-quality minority-class recognition.

### 3.4. Embedding Space Structure and Discriminative Behavior Analysis

To further investigate the underlying mechanisms driving the performance improvements of IRRL, we conduct an auxiliary analysis from two dimensions: representation structure and discriminative behavior, incorporating feature space visualizations, confusion matrices.

First, as observed from the t-SNE visualization results (Fig. 2 and Fig. 3), the full IRRL framework yields a more distinctly separable embedding distribution, wherein the minority-class samples exhibit more compact clustering within the feature space. In contrast, the baseline framework and several ablation variants suffer from varying degrees of class overlap and ambiguous decision boundaries. This indicates that IRRL more effectively enhances the stability of minority-class representations and augments inter-class separability during the representation learning process.

**Fig. 2:**
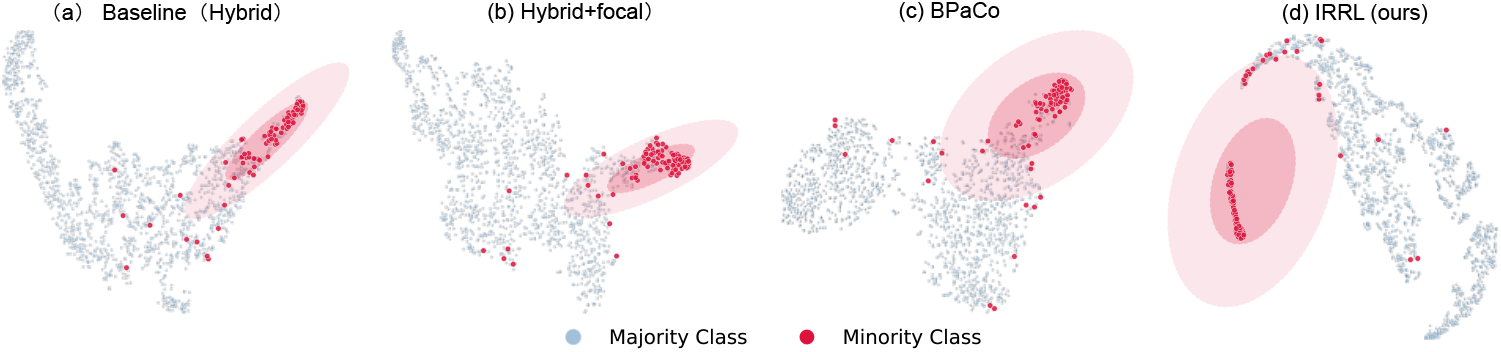
Shows the t-SNE visualization of feature distributions on the ISIC 2018 dataset. Different colors represent different classes. The visualization compares embedding structures learned by different methods, highlighting variations in inter-class separability and intra-class compactness under imbalanced learning conditions.

**Fig. 3:**
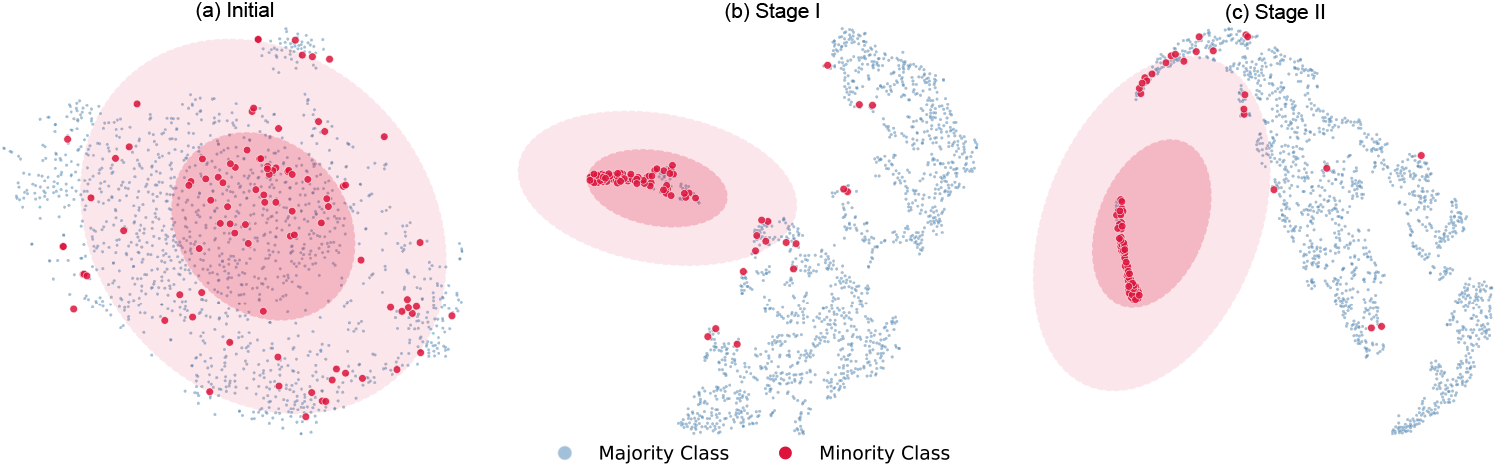
Illustrates the evolution of feature distributions across different training stages of IRRL. From initialization to Stage I and Stage II, feature representations gradually become more structured, with minority-class samples forming compact clusters and clearer decision boundaries emerging. This demonstrates the effectiveness of the proposed two-stage learning strategy.

The geometric metrics reported in Table 3 further corroborate these visual observations. Compared to the baseline model, IRRL exhibits a more consistent improvement in the Silhouette Score, presenting a more well-defined clustering structure across the overall embedding space. This suggests that the representation space learned by IRRL is highly conducive to the formation of clear-cut decision boundaries.

**Table 3:** Geometric analysis of embedding spaces, including intra-class variance, inter-class distance, and Silhouette Score.

| Method Category | Method / Training Stage | Intra-class Variance ↓ | Inter-class Distance ↑ | Silhouette Score ↑ |
| --- | --- | --- | --- | --- |
| Representative Methods | Baseline (Vanilla) | 0.0423 | 0.9034 | 0.6439 |
|  | Focal Loss | 0.0190 | 0.2380 | 0.7927 |
|  | BPaCo | 0.0088 | 0.3283 | 0.4868 |
| Proposed Method | (a) Initial State | 0.0051 | 0.0163 | 0.0353 |
|  | (b) Stage I | 0.0078 | 0.2696 | 0.8356 |
|  | (c) Stage II (IRRL) | 0.0131 | 0.3328 | 0.8402 |

Notably, the progressive geometric evolution reveals a continuous trajectory of improvement in the clustering quality of the feature spacefrom the initial state to Stage I, and subsequently to Stage II. This trajectory demonstrates that the two-stage training paradigm of IRRL”local representation refinement followed by global context modeling”yields substantial structural benefits in shaping a more discriminative embedding space.

To further validate the capability of IRRL to focus on minority-class lesion regions, we visualize the local implicit tokens (Fig. 4). The red bounding boxes delineate the critical lesion regions assigned high semantic confidence by the model during training. It can be observed that IRRL effectively highlights the local regions corresponding to minority-class lesions, whereas the influence of background or low-value areas is significantly suppressed. These visualizations offer intuitive evidence for the enhanced minority-class recognition performance reflected by quantitative metrics (such as the F1-score and MCC), thereby complementing the conclusions drawn from the t-SNE feature distribution analysis.

**Fig. 4:**
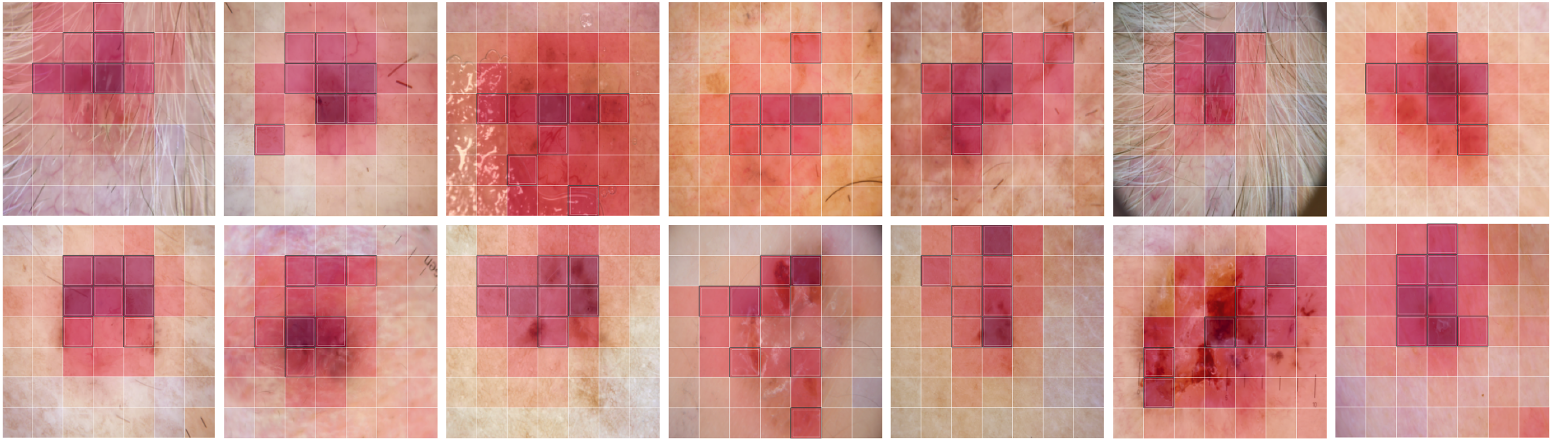
Presents the visualization of semantic confidence for local tokens. High-confidence regions correspond to lesion-relevant areas, indicating that the model effectively suppresses background responses while focusing on diagnostically meaningful regions, thereby improving interpretability and minority-class sensitivity.

Furthermore, as illustrated by the confusion matrices (Fig. 5), IRRL achieves more stable recognition of the minority class (BCC) while preserving robust discriminative performance on the majority class. This demonstrates that the proposed method does not elevate minority-class recall at the expense of overall classification consistency.

**Fig. 5:**
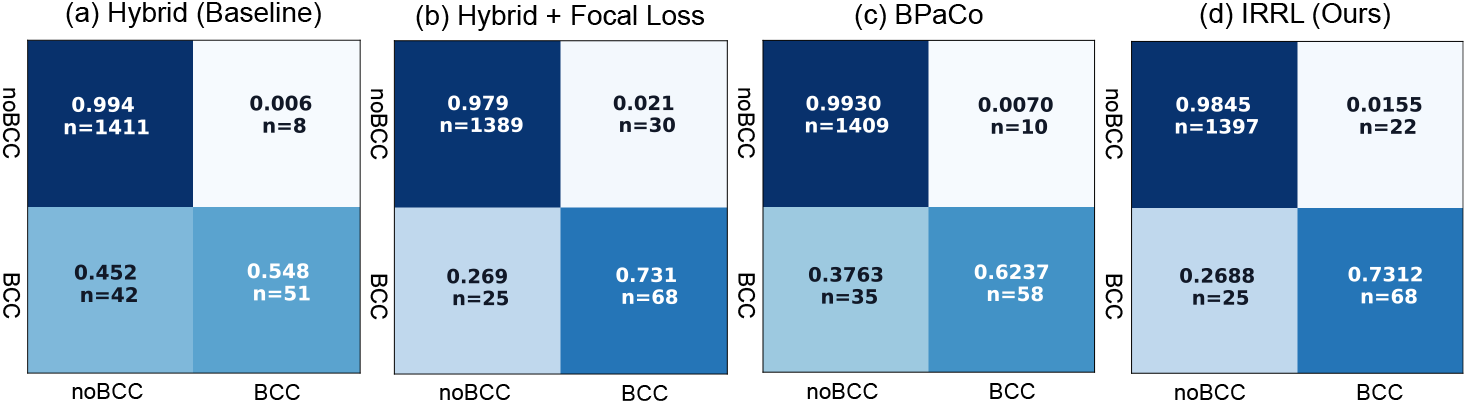
Shows confusion matrices of different methods on the ISIC 2018 dataset. Rows represent ground-truth labels and columns represent predicted labels. The comparison highlights differences in minority-class recognition, false positive suppression, and overall classification stability under imbalanced conditions.

In summary, the t-SNE visualizations, geometric embedding metrics, and confusion matrices provide consistent evidence from both qualitative and quantitative perspectives, collectively corroborating the mechanistic hypotheses proposed in this study. Specifically, during the representation learning phase, IRRL enhances the embedding quality of minority-class samples via semantic confidence modulation, difficulty-adaptive supervised contrastive learning, and minority-class prototype regularization. These refinements ultimately translate into superior class-imbalanced classification performance.

### 3.5. Cross-Dataset Results and Cross-Scenario Stability Analysis

To evaluate the applicability of IRRL across diverse medical image classification scenarios, we conduct supplementary experiments on three benchmark datasets: PAD-UFES-20, OCTID, and BUSI. Table 4 summarizes the AUC, F1-score, and MCC achieved by various methods across these datasets. The results demonstrate that IRRL consistently achieves superior overall performance across all three external datasets. Notably, IRRL attains an F1-score of 0.7904 and an MCC of 0.6711 on PAD-UFES-20; 0.8800 and 0.8735 on OC-TID; and 0.8941 and 0.8546 on BUSI, respectively, underscoring its robust cross-domain stability.

**Table 4:** Reports cross-dataset results on PAD-UFES-20, OCTID, and BUSI datasets. Metrics include AUC, F1-score, and MCC.

| Comparison Methods | PAD-UFES-20 (Skin) |  |  | OCTID (Retinal) |  |  | BUSI (Breast Ultrasound) |  |  |
| --- | --- | --- | --- | --- | --- | --- | --- | --- | --- |
|  | AUC | F1-score | MCC | AUC | F1-score | MCC | AUC | F1-score | MCC |
| Hybrid | 0.8998 | 0.7545 | 0.6147 | 0.9913 | 0.7097 | 0.7088 | 0.9465 | 0.8642 | 0.8177 |
| Hybrid + Focal | 0.8856 | 0.7527 | 0.5941 | **0.9965** | 0.8462 | 0.8397 | 0.9432 | 0.8642 | 0.8511 |
| ECL | 0.8710 | 0.7310 | 0.5720 | 0.9371 | 0.6250 | 0.6115 | 0.9150 | 0.7671 | 0.7118 |
| BPaCo | **0.9149** | 0.7866 | 0.6693 | 0.9869 | 0.7333 | 0.7310 | 0.9387 | 0.8250 | 0.7666 |
| **IRRL (Ours)** | 0.9085 | **0.7904** | **0.6711** | 0.9939 | **0.8800** | **0.8735** | **0.9559** | **0.8941** | **0.8546** |

The efficacy of IRRL is not contingent upon any specific dataset or single imaging modality. Given that PAD-UFES-20, OCTID, and BUSI correspond to smartphone clinical skin images, ophthalmic OCT scans, and breast ultrasound images, respectively, they exhibit substantial heterogeneities in acquisition devices, imaging principles, anatomical regions, and class distributions. Consequently, this set of experiments can comprehensively reflect the performance stability of the model across diverse medical imaging tasks. As demonstrated by the results in Table 4, IRRL consistently maintains high F1-scores and MCCs across multiple datasets. This indicates that its performance gains do not stem from localized overfitting to a single data distribution. Rather, it suggests that these improvements are driven by the effective refinement of the minority-class representation learning process under class-imbalanced conditions.

### 3.6. Ablation Study and Training Strategy Analysis

To validate the individual contributions of each constituent module to the overall performance, systematic ablation experiments are conducted on the ISIC 2018 dataset, with the results detailed in Table 5. Overall, the full IRRL model achieves the optimal F1-score and MCC among all variants, yielding 0.7432 and 0.7267, respectively, which highlights a pronounced synergistic complementarity among the constituent modules.

**Table 5:** Ablation study of IRRL on the ISIC 2018 dataset, evaluating the contribution of different components and training strategies. Performance is reported in terms of AUC, Sensitivity, Specificity, F1-score, and MCC.

| Group | Configuration | AUC | Sens. | Spec. | F1-score | MCC |
| --- | --- | --- | --- | --- | --- | --- |
| Feature Level | Global Image-level (w/o Implicit Tokens) | 0.9675 | 0.6344 | 0.9852 | 0.6821 | 0.6650 |
| Module Ablation | w/o Contrastive Loss ($\mathcal{L}_{DASC}$) | 0.9751 | 0.6344 | 0.9782 | 0.6448 | 0.6220 |
| | w/o Prototype Regularization ($\mathcal{L}_{proto}$) | 0.9582 | 0.6774 | **0.9887** | 0.7326 | 0.7192 |
|  | w/o Semantic Confidence | 0.9762 | 0.6559 | **0.9887** | 0.7176 | 0.7045 |
|  | w/o Difficulty Adaptation | 0.9739 | 0.5914 | 0.9873 | 0.6627 | 0.6486 |
|  | w/o Class-Imbalance Boost | 0.9726 | 0.6667 | 0.9796 | 0.6739 | 0.6528 |
| Training Strategy | Direct End-to-End | 0.9754 | 0.6989 | 0.9831 | 0.7143 | 0.6962 |
|  | Stage I Only | 0.9603 | **0.7742** | 0.9753 | 0.7200 | 0.7022 |
|  | Synchronous Joint Training | **0.9778** | 0.6344 | 0.9866 | 0.6901 | 0.6746 |
| Proposed | **Full IRRL Model** | 0.9702 | 0.7312 | 0.9845 | **0.7432** | **0.7267** |

In terms of specific module contributions, removing the supervised contrastive learning term results in the most pronounced performance degradation. This indicates that supervised contrastive learning plays a pivotal role in enhancing the discriminability of local representations and improving decision boundary modeling for minority classes. Upon the ablation of the difficulty-adaptive mechanism, the F1-score and MCC also exhibit notable deterioration. This demonstrates that under class-imbalanced conditions, explicitly elevating the training contributions of hard examples and critical local regions is of great significance for intensifying minority-class learning. Removing the semantic confidence module similarly leads to a performance drop, implying that modeling the semantic importance of local regions facilitates the suppression of background noise and enhances the representation quality of effective lesion areas. After omitting the prototype constraint, although the degradation in classification performance is relatively mild, a combination with geometric metrics reveals an increase in intra-class variance. This suggests that the prototype constraint acts more on augmenting the internal structural stability of the minority classes, rather than merely operating on the classifier output layer.

In addition to module ablation, this study also compares the efficacy of different training strategies. The results reveal that direct end-to-end training, retaining only Stage I, or synchronous joint training all fail to attain the performance level of the full IRRL framework. Instead, the two-stage training paradigmemploying “local representation optimization first, followed by global modeling via a fine-tuning strategy in Stage II”yields the optimal performance. This outcome illustrates that the relationship between local representation optimization and global context modeling is not a naïve superposition, but rather requires a rational training organization to achieve synergy. Prioritizing the enhancement of token representation quality at the local level before conducting image-level discriminative learning at the global level is highly conducive to exploiting the complementary strengths of the CNN and Transformer, thereby yielding more robust final results.

Synthesizing the ablation results and the embedding space analysis, it becomes further evident that the performance gains of IRRL do not originate from the incidental gains of a single module. Instead, they are derived from the collective interplay of receptive-field-based local embedding modeling, semantic confidence modulation, difficulty-adaptive optimization, minority-class prototype constraints, and the two-stage training paradigm. Such a multi-tier, complementary design empowers IRRL to effectively refine the stability and discriminability of minority-class representations under class-imbalanced conditions, without altering the original data distribution.

### 3.7. Complexity and Performance Trade-off Analysis

In addition to classification performance, we further compare the training memory consumption across different methods. As illustrated in Table 6, compared to the baseline Hybrid framework, the training memory consumption of IRRL only marginally increases from 1.916 GB to 2.115 GB, corresponding to a modest additional overhead of a mere 10.4%. Concurrently, the F1-score and MCC are elevated to 0.7432 and 0.7267, respectively.

**Table 6:**
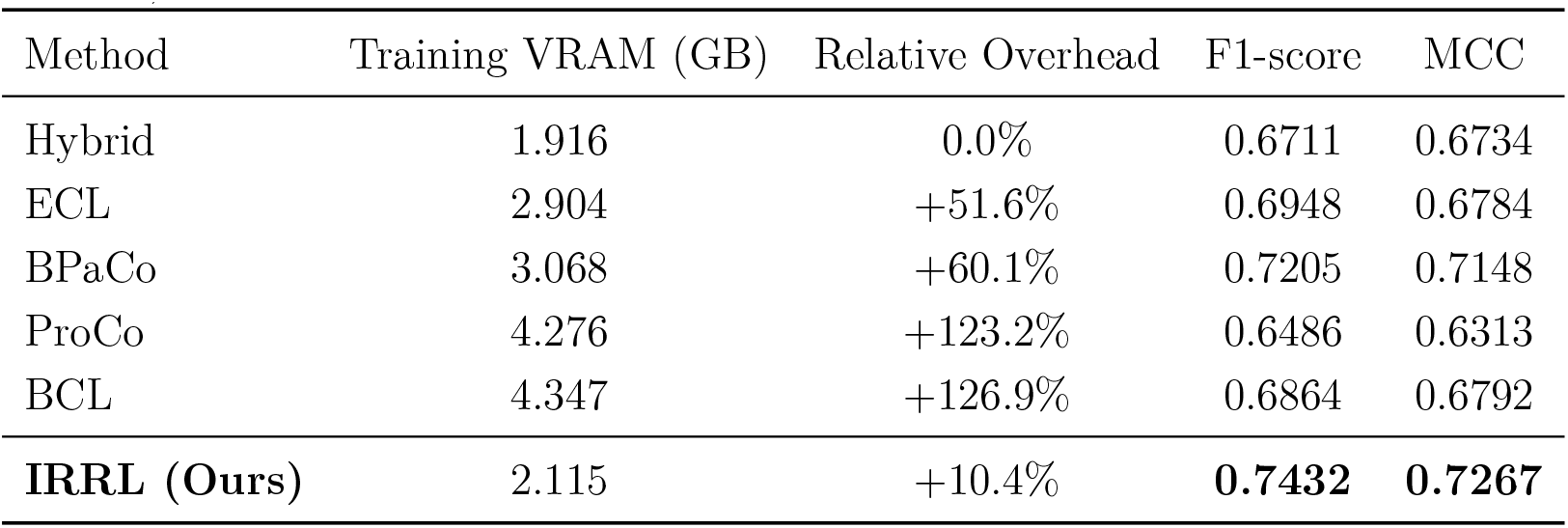
Comparison between IRRL and representative methods in terms of computational cost and classification performance. Metrics include training VRAM, relative overhead, F1-score, and MCC.

In contrast, several competing methods yield only marginal performance gains while incurring significantly higher memory costs. These results demonstrate that IRRL strikes a favorable balance between performance enhancement and computational expense. The overall performance improvements are not primarily attained by drastically inflating model complexity, but rather stem from the effective optimization of the representation learning process.

## 4. Discussion and Conclusion

This study explores class-imbalance mitigation from the perspective of representation-level refinement rather than relying solely on classifier-level compensation. The experimental results suggest that improving minority-class representations before global classification can provide a more balanced trade-off among different aspects of classification performance.

In extremely imbalanced scenarios, the majority class tends to dominate the direction of gradient updates, leading to the compression of the feature space for the minority class. Under such highly challenging conditions, IRRL achieves relatively stable performance, which can be attributed to its capacity to facilitate the structural decoupling and dynamic reallocation of the feature space. First, the receptive-field-based implicit token construction preserves crucial spatial topological priors inherent in medical images, while concurrently mitigating the contextual fragmentation induced by explicit patch division. Building upon this, the semantic confidence and difficulty-adaptive mechanisms forge a dual-gating strategy: the former suppresses low-value background noise in the spatial dimension, whereas the latter amplifies the gradient contributions from hard and high-risk samples in the instance dimension. Coupled with the minority-class prototype regularization that constrains intra-class variance, IRRL mitigates the tendency of minority-class representations to be overwhelmed or diluted by majority-class patterns within the deep feature space, thereby establishing a more reliable representational foundation for learning discriminative decision boundaries.

Despite demonstrating highly satisfactory performance under multi-modality settings, IRRL still presents several limitations. First, the semantic confidence estimation of local lesion regions is, to some extent, contingent upon the quality of intermediate representations. When confronting challenging cases such as severe occlusions, dense artifacts, or advanced diffuse lesions, the confidence estimation module may exhibit fluctuations, which could potentially introduce sub-optimal optimization signals. Secondly, the improvement in the embedding space structure is predominantly corroborated by empirical geometric analyses and visualizations, lacking a rigorous theoretical characterization of the generalization behavior of the difficulty-adaptive contrastive loss under long-tailed distributions. Furthermore, the current framework does not explicitly incorporate adversarial domain adaptation mechanisms; consequently, its robustness under severe clinical domain shifts remains to be validated on larger and more diverse independent cohorts.

The current experimental setup primarily focuses on binary classification tasks aimed at differentiating between clinically relevant minority and majority classes. However, real-world clinical data frequently exhibit more complex long-tailed multi-class distributions. Extending IRRL to such scenarios will necessitate generalizing the current single minority-class prototype constraint into a dynamic multi-prototype geometric structure, so as to resolve the feature entanglement among multiple tail classes. Future work will further explore robust modeling within the context of multi-class long-tailed medical image classification, with a particular focus on dynamic prototype learning and the enhancement of generalization capabilities under complex clinical domain shifts.

## Data Availability

The datasets analyzed in this study are publicly available from their respective repositories. The ISIC 2018, PAD-UFES-20, OCTID, and BUSI datasets can be accessed through the links provided in the Data Availability Links section and the corresponding references cited in the manuscript.

https://challenge.isic-archive.com/data/

https://data.mendeley.com/datasets/zr7vgbcyr2/1

https://dataverse.scholarsportal.info/dataverse/OCTID

https://www.kaggle.com/datasets/aryashah2k/breast-ultrasound-images-dataset

## CRediT authorship contribution statement

**Manyuan Cheng:** Conceptualization, Methodology, Software, Validation, Formal analysis, Investigation, Data curation, Visualization, Writing – original draft. **Cuiyin Liu:** Conceptualization, Methodology, Supervision, Project administration, Writing – review & editing. **Lei Gu:** Methodology, Supervision, Writing – review & editing.

## Declaration of competing interest

The authors declare that they have no known competing financial interests or personal relationships that could have appeared to influence the work reported in this paper.

## Funding

This work was supported by the National Natural Science Foundation of China (Grant No. 11773012).

## Data availability

The datasets analyzed in this study are publicly available from their respective sources. The ISIC 2018, PAD-UFES-20, OCTID, and BUSI datasets can be accessed through the publicly available sources cited in the manuscript.

## Notes

### Competing Interest Statement

The authors have declared no competing interest.

### Author Declarations

This study used only openly available, de-identified human medical imaging datasets that were publicly accessible before the initiation of the study. The datasets were obtained from the following public sources: ISIC 2018: https://challenge.isic-archive.com/data/ PAD-UFES-20: https://data.mendeley.com/datasets/zr7vgbcyr2/1 OCTID: https://dataverse.scholarsportal.info/dataverse/OCTID BUSI: https://www.kaggle.com/datasets/aryashah2k/breast-ultrasound-images-dataset No new participants were recruited, and no new human data or personally identifiable information were collected by the authors.

